# Longitudinal genomic surveillance of multidrug-resistant *Escherichia coli* carriage in critical care patients

**DOI:** 10.1101/2023.08.12.23293895

**Authors:** Mira El Chaar, Yaralynn Khoury, Gavin M. Douglas, Samir El Kazzi, Tamima Jisr, Shatha Soussi, Georgi Merhi, Rima A. Moghnieh, B. Jesse Shapiro

## Abstract

**Background:** Colonization with multidrug resistant *E. coli* strains causes a substantial health burden in hospitalized patients. We performed a longitudinal genomics study to investigate the colonization of resistant *E. coli* strains in critically ill patients, and to identify evolutionary changes and strain replacement events within patients.

**Methods:** Patients were admitted to the intensive care unit and haematology wards at a major hospital in Lebanon. Perianal swabs were collected from participants on admission and during hospitalization, which were screened for extended-spectrum beta-lactamases and carbapenem-resistant Enterobacterales. We performed whole-genome sequencing and analysis on *E. coli* strains isolated from patients at multiple time points.

**Results:** The *E. coli* isolates were genetically diverse, with 13 sequence types (STs) identified among 21 isolates sequenced. Five patients were colonized by ST131 encoding CTX-M-27, a type of beta-lactamase gene not previously been observed in Lebanon. Among the eight patients whose resident *E. coli* strains were tracked over time, five harbored the same *E. coli* strain with relatively few mutations over the 5 to 10 days of hospitalization. The other three patients were colonized by different *E. coli* strains over time.

**Conclusion:** Our study provides evidence of strain diversity within patients during their hospitalization. While strains varied in their antimicrobial resistance profiles, the number of resistance genes did not increase over time. We also show that ST131 encoding CTX-M-27, which appears to be emerging as a globally important multidrug resistant *E. coli* strain, is also prevalent among critical care patients and deserves further monitoring.

## 1. Introduction

Multidrug-resistant Gram-negative bacteria represent an important threat in hospital settings (1–3). During hospitalization, patients may be colonized with bacteria that develop resistance due to changes in gene expression, or that evolve resistance via point mutation or horizontal gene transfer (4–6). In hospitalized patients, resistant strains’ survival and replication may depend on the selection pressure exerted by antibiotics (6). When resistant bacteria colonize the gastrointestinal tract, this provides an opportunity for transfer of resistance genes among pathogens and gut microbiome commensal bacteria (7–9). Multidrug-resistant *Escherichia coli* are a burden on healthcare systems, and are often responsible for treatment failures in patients (10–12). However, the success of the globally prevalent *Escherichia coli* sequence type 131 (ST131) is not easily explained simply by its antibiotic resistance profile and likely involves diverse colonization and virulence factors (13, 14). Resistant bacteria disseminate among critically-ill patients in hospitals, causing life-threatening infections (7, 15). The pathogenicity of *E. coli* is multifaceted, including genetic and environment factors allowing *E. coli* to expand its range of infection beyond the intestine, which increases disease severity (16–19).

Mixed infections of distinct *E. coli* strains within a single patient have been observed previously, and these strains can also evolve within patients (20). Studies tracking *E. coli* diversity within patients over time are still relatively rare and have focused mainly on extraintestinal infections. For example, the transition of *E. coli* from urine to blood usually involves very few genetic changes, but occasionally involves colonization by genetically distant strains (21). In acute infections, extraintestinal *E. coli* can rapidly evolve hypermutator phenotypes, generating dozens to hundreds of mutations that could provide adaptation to new tissue types (22). Over time scales of a few years, up to 32% of samples U.S. military personnel were colonized by multiple distinct *E. coli* phylogroups, indicating co-existence of distinct strains or replacement of one strain by another in the gut (23). In a UK hospital study, over 25% of patients were colonized by multiple distinct strains (19). Whether such strain dynamics occur during intestinal colonization over shorter time scales, or within hospitalized patients, remains unclear. We hypothesize that the hospital environment provides a diverse pool of strains for co-colonization or strain replacements, and may also select for antibiotic resistance genes as critically ill patients are treated with broad-spectrum antibiotics.

In Lebanon, the prevalence of extended-spectrum beta-lactamase (ESBL) producing *E. coli* has increased over time, from 12% in 2005 to 25% in 2012 (24). *E. coli* was the most frequently isolated bacterium from bloodstream infections (45.6%), with 79.6% of the isolates producing ESBLs (25). Most studies to date have described the prevalence of resistant *E. coli* strains isolated from hospitals or community settings in Lebanon (26–33). Others described their prevalence in animals and the environment (22, 34–42). Here, we analysed whole genome sequence data using a longitudinal approach to describe the colonization and evolution of multidrug resistant *E. coli* in critically ill patients in Lebanon.

## 2. Methods

### 2.1. Study design

A prospective observational cohort study was conducted over a six-month period from June 2021 to December 2021 at Makassed General Hospital in Lebanon. This is a 200 bed hospital located in a heavily populated area of Beirut with a medically underserved population. It serves nearly 15,000 inpatients per year that are mostly from middle to low socioeconomic status groups.

### 2.2. Sampling, bacterial culture and data collection

A standard protocol for infection and prevention control to screen for any patient admitted to the intensive care unit and to the hematology oncology unit has previously been established at Makassed General Hospital. A perianal swab was collected from each patient and screened for extended-spectrum beta-lactamases and carbapenem-resistant Enterobacterales (ESBL and CRE, respectively). In total, 144 patients were admitted to the intensive care and oncology units; they were recruited for this study and followed during their hospitalization. Only patients that were hospitalized for more than three days were included. In total, 97 were dropped because of their early discharge, death, or transfer to another hospital ward, leaving 47 remaining patients. Perianal swabs were collected by healthcare workers at admission and during hospitalization. A minimum of five days was required between the first, second or third collection time. A standardized questionnaire for each patient was filled by the resident consultee. The questionnaire included information about age, gender, cause of hospitalization, duration of hospitalization, antibiotics exposure (past and during hospitalization), types of antibiotics used, and bacterial infection during hospitalization.

### 2.3. Ethical consideration

This study was approved by the institutional review board of Makassed General Hospital. The ethical committee reviewed and approved the study protocol. An informed consent was provided by the participants that have a decision-making capacity. Participants lacking capacity were enrolled following discussion with their relatives that are involved in their care.

### 2.4. Bacterial Culture

Anal swabs were cultured on three different culture media plates: MacConkey agar without antibiotics, MacConkey agar with ertapenem (0.5 mg/L) to isolate carbapenamase-producing bacteria, and MacConkey agar with ceftriaxone (4 μg/ml) for isolation of ESBL-producing bacteria. All plates were incubated at 37 °C for 24 h. Following bacterial growth, two colonies of the same color, morphology and shape were picked and pooled, then taxonomically classified using matrix-assisted laser desorption/ionization time-of-flight mass spectrometry (MALDI-TOF). Antimicrobial susceptibility using the disk diffusion method was determined for all *E. coli* isolates for the following antibiotics: amoxicillin (20 μg), amoxicillin/clavulanic acid (20/10 μg), cefepime (30 μg), ceftriaxone (30 μg), piperacillin-tazobactam (100/10 µg), ceftazidime-avibactam (20/10 μg), cephalothin (30 μg), ertapenem (10 μg), fosfomycin (200 μg), trimethoprim/sulfamethoxazole (1.25/23.75 μg), ciprofloxacin (5 μg), colistin (10μg), imipenem (10 μg), nitrofurantoin (300 μg), amikacin (30 μg), tetracyclin (30 μg), and gentamicin (10 µg).

### 2.5. Whole genome sequencing

Genomic DNA was extracted from the *E. coli* isolates using QIAamp DNA Mini kit (Qiagen) following the manufacturer’s guidelines. DNA concentrations were measured using Qubit (Thermo Fisher Scientific) with the Qubit dsDNA HS Assay Kit (Thermo Fisher Scientific). Sequencing libraries were made with Nextera XT DNA Library Preparation Kit (Illumina) according to manufacturer’s instructions. Illumina sequencing was performed on a Novaseq- 6000 producing paired-end 2 × 150 bp reads, with an average base quality (Phred score) of 36 and G + C content of 54.1%.

### 2.6. Genomic Analysis

Reads were processed with Trimmomatic v0.39 (43) to remove adaptor sequences and quality filter the reads; we specified the TruSeq3 adapters and the quality filtering parameters of LEADING:3, TRAILING:3, SLIDINGWINDOW:4:15, and MINLEN:30. The assembly was performed using SPAdes Genome Assembler v3.15.4 with default parameters (44). Assemblies were annotated with Prokka v1.14.5 (45) with the kingdom specified as bacteria. The resulting annotated genes were processed by Panaroo v1.3.0 to create the core genome alignment and pangenome (46). This was run with the ‘strict’ clean mode parameter and with the ‘clustal’ option specified for the core genome alignment. Plots were generated using Roary v3.12.0 including the pangenome frequency plot, a presence and absence matrix against a tree and a pie chart of the pangenome, breaking down the core, soft core, shell and cloud (47). The phylogenetic tree along with the pangenome of the isolates were visualized with Phandango v1.1.0 (48). A maximum-likelihood phylogeny was generated based on the core genome alignment from Panaroo input into RaxML (46, 49). The resulting tree was visualised with the Interactive Tree of Life (iTOL v6) (50). SRST2 (v0.2.0) was used to identify MLST types and serotype (51). Phylogroups were classified based on the ClermonTyping method (52). FastANI v1.33 was used to calculate the Average Nucleotide Identity (ANI) of orthologous genomic regions between pairs of genome assemblies (53). The presence of antimicrobial resistance genes, putative virulence factors, and plasmid replicons were studied using ABRicate with the ResFinder database, Virulence Factor database, and PlasmidFinder database (54). We used Snippy v4.6.0 to compare *E. coli* genomes from the same patient and identify single nucleotide variants (SNVs). The reference genome used was the *E. coli* strain identified at T0 (at hospital admission) or at T1 (the first collection time during hospitalization) (55). The major allele frequency distribution was also based on the identified variants, including SNVs and insertions/deletions.

### 2.7. Data Availability

All sequence data has been deposited in DDBJ/ENA/GenBank under BioProject PRJNA962847 and accession numbers SAMN35447827, SAMN35449847, SAMN35450267, SAMN35690035, SAMN35690036, SAMN35690056, SAMN35690090, SAMN35690097, SAMN35690110, SAMN35709899, SAMN35709920, SAMN35711041, SAMN35711058, SAMN35711154, SAMN35713710, SAMN35714296, SAMN35716244, SAMN35728356, SAMN35728381, SAMN35728629, SAMN35731198 and SAMN35731315.

## 3. Results

### 3.1. Study participants and microbial carriage

In total, 20 (43%) patients were eligible for this study, all of whom were colonized by antibiotic resistant isolates. Throughout their hospitalization, 14 (70%) patients were colonized with *E. coli*, one with *Raoultella ornithinolytica* and one with *Klebsiella pneumoniae*. Four patients were colonized by multiple bacterial species during their hospitalization. Here, we focus on the *E. coli*-colonized patients. The median age of patients infected with *E. coli* was 67 years (range = 17 - 88 years), eight were female (57%) and six (43%) were male. These 14 patients were admitted to different hospital wards: the intensive care unit (N=7), the haematology and oncology ward (N=3) and the isolation ward (N=4). The median duration of hospitalization for these patients was 29 days (range = 9 - 73 days). The cause of admission was either pneumonia, chemotherapy, bleeding, or sepsis (**Table S1**). In total, 11 out of 14 patients had an indwelling catheter during hospitalization and 13 patients were under corticosteroid therapy. The patients were exposed mostly to broad-spectrum antibiotics during their hospitalization, including: meropenem which was used in 50% of patients; colistin and piperacillin/tazobactam in 50%; amikacin, ceftriaxone and ceftazidime/avibactam in 50%; and levofloxacin in 29% of patients. Vancomycin was used in 43% of patients; trimethoprim-sulfamethoxazole, teicoplanin, gentamycin and clarithromycin were used in less than 20% of patients. The median time between first sample collection (T0) and second collection (T1) was 9 days (range = 5 - 28 days). During hospitalization, 7/14 (50%) of patients were documented to have infectious episodes (Table S1), including respiratory tract infections (N=5), followed by bacteremia (N=5), urinary tract (N=3) and skin and soft tissue (N=3).

### 3.2. Genomic typing and phylogeny of *E. coli* isolates

In total, 12 out of 14 patients infected with *E. coli* were selected for further analysis, of which eight patients had isolates from multiple time points, yielding 22 whole-genome sequences in total. Two patients (MGH 3A/B and MGH 12A) and two isolates (MGH 2A and MGH 6B) were dropped since their assembly sizes exceeded 8 Mbp, much larger than expected for an *E. coli* genome. Two of these assemblies also had median SNV major allele frequencies in the 0.93-0.97 range, whereas all others were very close to 1 (**Figure S1**). This suggests that some of the unexpectedly large assemblies could have been due to contamination, or due to pooling colonies of two distinct *E. coli* strains. The remaining 22 genomes considered for further analysis can be considered single strains.

These 22 *E. coli* genomes were genetically diverse, with 13 STs (ST10, ST44, ST69, ST131, ST224, ST227, ST 540, ST648, ST1286, ST1431, ST1491) identified (**Figure 1**). The most common was ST131 which was carried by 5/12 patients (41%).

**Figure 1:**
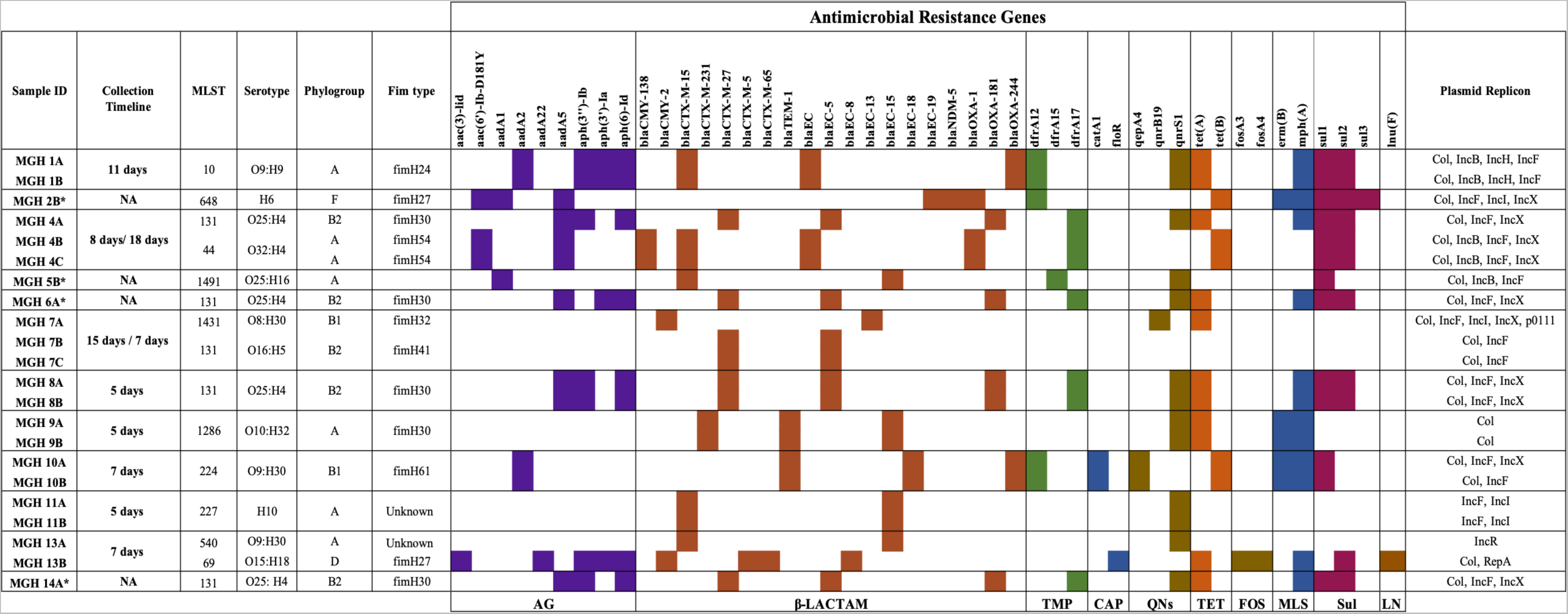
Distribution of antimicrobial resistance genes across the *E. coli* genomes recovered from patients. Genetic determinants of resistance are grouped according to their corresponding antimicrobial classes, which are colour coded. Sequence type profiles, serotypes, phylogroup, and Fim type are indicated for each isolate. The time differences between T0/T1 or T1/T2 are indicated under Collection Timeline. *MGH 2B, MGH 5B, MGH 6A and MGH 14A strains were sequenced only at T0 or T1. The plasmid replicons identified in each isolate were included in the table. The patients are differentiated by numbers and each letter next to the number represent the different time points of collection: A (T0), B (T1) and C (T2)

We inferred the maximum-likelihood phylogeny of these isolates using a core-gene alignment. We included *E. coli* genomes previously sequenced from clinical samples collected in Lebanon (31, 32). Based on this phylogeny, we identified 5 phylogroups. In total, five isolates were phylogroup C and eight were phylogroup A. The ST131 strains (within phylogroup B2) were the most frequent (N=7). Phylogroups F and D were each represented by one isolate. In total, 11 serotypes were identified with the most common being O25:H4 and O9:H30 (**Figure 2**).

**Figure 2:**
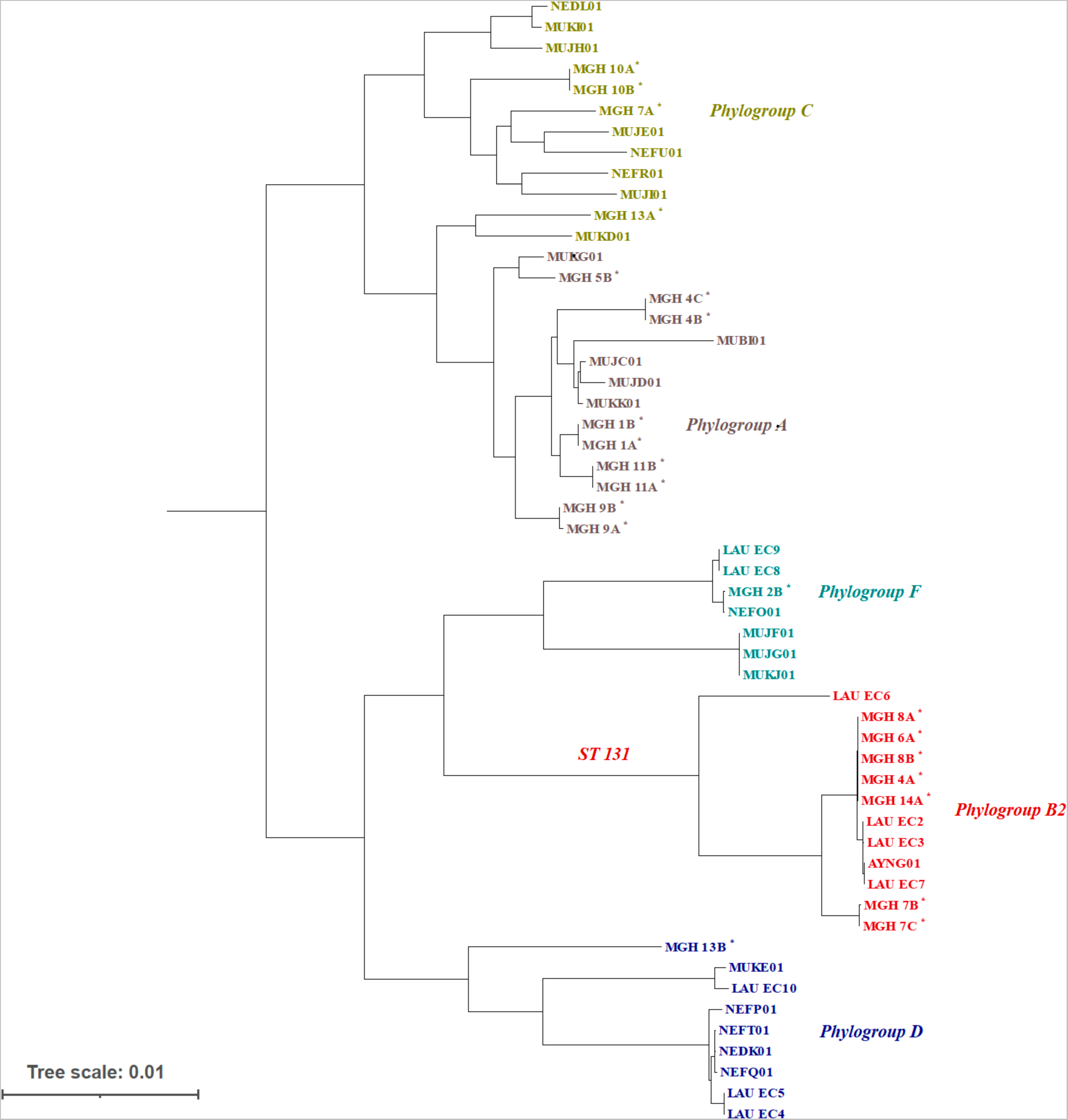
Maximum likelihood phylogenetic tree of 54 *E. coli* isolates from clinical samples in Lebanon. The tree includes genomes sequenced in the current study (indicated by asterisks). The phylogroup of the samples are shown in different colors.

Of the six patients sampled at two time points 5-11 days apart, five patients (MGH 1, MGH 8, MGH 9, MGH 10, MGH 11) had pairs of isolates that clustered very closely on the phylogeny (**Figure 2**) and had identical profiles of AMR genes (**Figure 1**). This is consistent with *E. coli* lineages persisting in patients between sampling points, although we cannot exclude transmission events of near-identical genomes among patients; for example, the two isolates from patient MGH 8 were very closely related to isolates from patients MGH 4, 6, and 14, suggesting possibly recent transmission of ST131 strains. In contrast, the two isolates from patient MGH 13 sampled seven days apart came from entirely different phylogroups (C and D; **Figure 2**) and had distinct AMR gene and plasmid profiles (**Figure 1**). This suggests either a mixed infection in patient MGH 13 (with phylogroup C sampled first and D sampled second), or a strain replacement that occurred between time points.

### 3.3. Prevalence and pangenome variation of ST 131 strains

Among the 12 patients with sequenced isolates, five harbored the ST131 type. Of these, four strains belonged to the O25:H4 serotype and fimH 30 variant and one strain to the O16:H5 serotype and fimH 41 variant. ST131 formed a distinct clade on the phylogenetic tree, classified as phylogroup B2 (**Figure 2**). All ST131 genomes encode *bla*_CTX-M-27_, bla_EC-5_, and the IncF plasmid, and 80% of them also encode *bla*_OXA-181_. They included type 1 Fimbriae, P fimbriae, and enterotoxin. All ST131 genomes were highly genetically similar, with average nucleotide identity values ranging from 99.5% to 99.9% (**Figure S2**). A pangenome analysis revealed 3308 core genes present in all isolates, 85 ‘soft core’ genes, 2520 ‘shell genes,’ and 2653 ‘cloud’ genes. The gene presence/absence patterns are shown alongside the phylogeny and strain typing information (**Figure 3**). Notably, ST131 strains with the O25:H4 serotype have a unique pattern of gene presence/absence, which is distinct from other ST131 strains and from other phylogroups.

**Figure 3:**
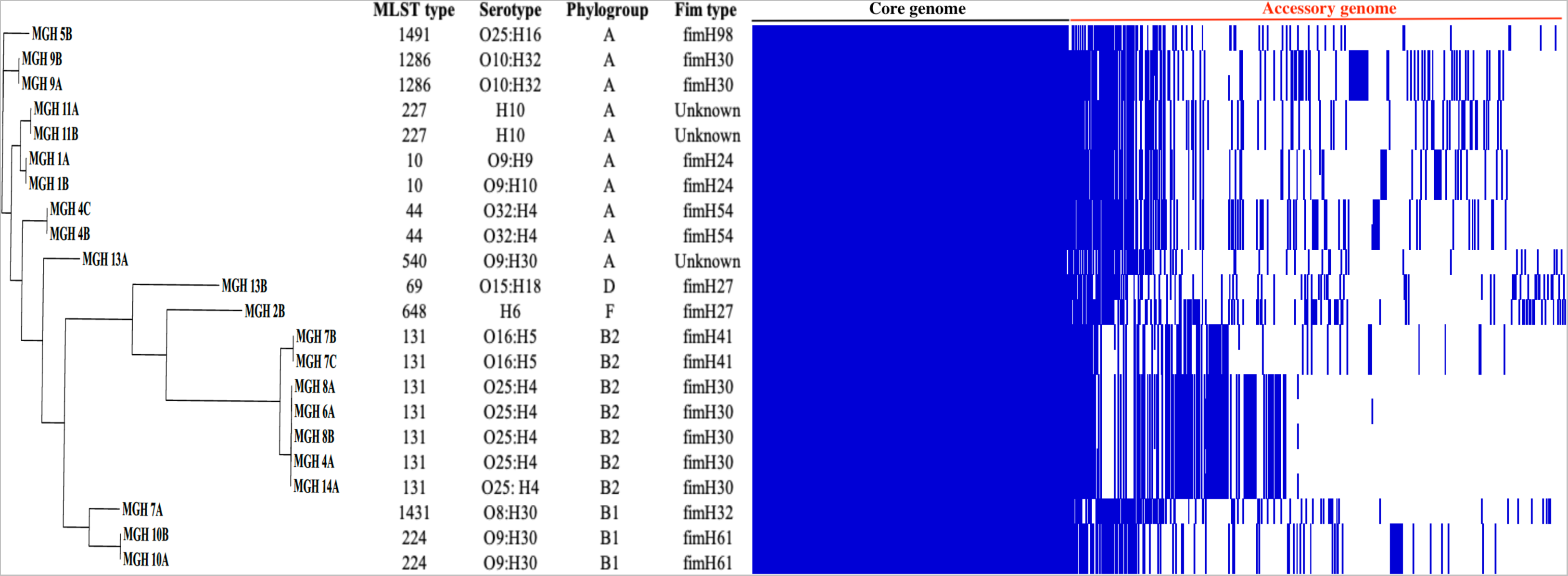
Gene presence/absence matrix from pangenome analysis of 22 *E. coli* isolates. The Pangenome and maximum likelihood tree and distribution of accessory genes were visualized using Phandango. Blue and white represent the presence and absence of genes, respectively.

### 3.4. Antimicrobial susceptibility profiles and genetic determinants of AMR

Among the 14 patients carrying *E. coli*, 13 carried multidrug-resistant (MDR) strains. Only one patient (MGH 11) carried a strain that was resistant to cephalosporins, yet it was susceptible to all other antibiotics (**Table S2**).

Susceptibility testing was performed on 29 *E. coli* isolates collected from patients at different time points. Identical strains (defined as ANI of 99.9% or more) isolated from the same patient at different time points were omitted from the analysis (N=7). Among the remaining 22 *E. coli* strains, 55% (N=12/22) were ESBL, 5% (N=1/22) CRE, and 41% (N=9/22) ESBL/CRE. All patients enrolled in this study carried resistant strains upon admission. A total of 95% (N=21/22) were resistant or intermediate to cefotaxime, 86% to ceftriaxone (N=19/22), 54% (N=12/22) to ceftazidime, 63% (N=14/22) to ciprofloxacin, 50% (N=11/22) to ertapenem, 32% (N=7/22) to imipenem and meropenem, and 10% (N=2/22) to ceftazidime/avibactam (**Table S2**).

The genomes of these isolates were screened for known genetic determinants of AMR using the ABRicate pipeline. More than 46 different acquired AMR genes were detected; β-lactamases (N=19 genes) and aminoglycosidases (N=9 genes) were among the most common, and patients contained 2-16 resistance genes, with a median of 11 per patient (**Figure 1**). Among the 12 patients with non-identical *E. coli* strains, Class A, C and D beta-lactamases were mostly identified, including *bla*_CTX-M-15_ (N=5/12, 41%), *bla*_CTX-M-27_ (N=5/12, 41%), *bla*_EC-5_ (N=5/12, 41%), *bla*_EC-15_ (N=4/12, 33%), *bla*_OXA-181_(N=4/12, 33%). The beta-lactamases *bla*_NDM-5_ and *bla*_OXA-244_ were seen in one and two patients respectively. Several other AMR genes were also detected (**Figure 2**), with *qnrS1* (encoding quinolone resistance) being quite prevalent (N=9/12, 75%).

### 3.5. Plasmid and virulence factor identification

The ABRicate pipeline was also used to identify the incompatibility groups for the recovered plasmid replicons (**Figure S3**). IncFI was the most common plasmid (73%; N= 16/22), followed by IncFII (63%; M= 14/22), IncX (41%, N= 9/22), IncI (23%; N= 5/22), IncB (27%; N=6/22) and IncH (14%; N= 3/22).

We screened the isolates for virulence genes which are often encoded by *E. coli*, and detected a wide variety (**Figure S4**). Some virulence genes were commonly detected, including *ompA* (100%) which is an outer membrane protein required for conjugation. Iron regulatory proteins were detected in 86% (N= 19/22) of isolates.

We recovered some of the known markers of intestinal and extraintestinal virulence in some isolates, such as the invasion and evasion factors (*kpsM* and *kpsD* [41%, N= 9/22]) and adherence factors (*fim* [86%, N= 19/22] and *pap* genes [41%, N= 9/22]) that are associated with intestinal and extraintestinal infections in humans. One patient (MGH 9) harboured an enteroaggregative *E. coli* strain with an anti-aggregation gene, and another patient (MGH 5) an enterotoxigenic *E. coli* with heat-labile (LT) and heat-stable (ST) enterotoxins genes. Other virulence genes detected in the study included *chu*, which codes for an outer membrane hemin receptor (41%, N= 9/22), and the enterotoxin gene, *senB* (32%, N= 7/22).

### 3.6. Strain dynamics within patients

A comparative genomic analysis was performed to compare the strains isolated from patients at different time points. Among the eight patients who were followed over time, five (MGH 1, MGH 8, MGH 9, MGH 10 and MGH 11) had very closely-related *E. coli* isolates at T0 and T1 (**Figure 1**, **Figure 2**). The duration of hospitalization for these patients ranged between 5 and 11 days, and genomes differed by an average of 16 SNVs (range 10-22 SNVs). In contrast, three patients (MGH 4, MGH 7 and MGH 13) were colonized by a highly divergent strain (different by > 1000 SNVs or <97% ANI; **Figure S2**) between time points, leading to marked changes in AMR gene profiles (**Figure 1**). In two patients, (MGH 4 and MGH 7), there was a putative strain replacement between T0 and T1, with the same strain persisting between T1 and T2, differing by a few SNVs (**Figure 4**). Comparing only closely-related pairs of genomes from the same patient yielded no evidence for a molecular clock (Spearman’s correlation between number of SNVs and number of days separating isolates, rho = 0.24, *p* = 0.60). The lack of clock signal could be due to a small sample size, short duration of sampling, or due to sampling only one isolate per patient per time point.

**Figure 4:**
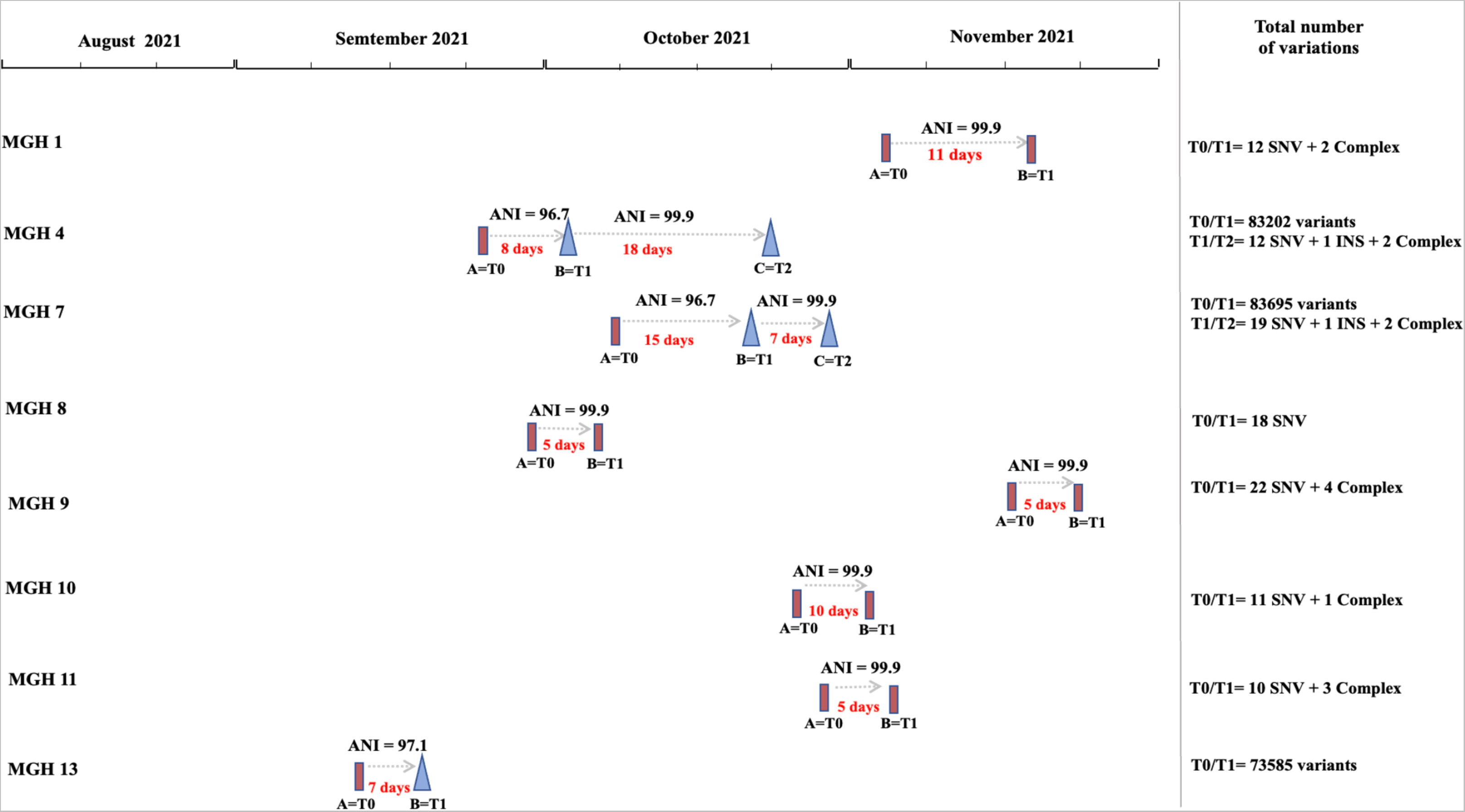
Summary of *E. coli* genetic diversity and putative strain replacements within patients over time. Each patient is shown on a separate row, with time indicated along the horizontal axis. *E. coli* genomes are indicated with rectangles or triangles, connected by arrows showing the number of days between samples and their average nucleotide identity (ANI). Genomes sharing a high ANI (99.9% or more) are shown with the same shape, and those with lower ANI (different strains) are shown in different shapes. The total number of genetic variations within each patient are indicated on the right: INS (insertion sequences), and complex variants (the combination of SNVs and multiple nucleotide polymorphism).

Of the 120 genetic variants (SNVs or complex variants of multiple mismatches nucleotides or insertions/deletions) observed between closely-related isolates from the same patient, half (N=60/120) were located in non-coding regions and 29% (N= 35/120) were synonymous changes. Most occurred in hypothetical proteins (N=18/35, 51%) (**Table S3)**. Missense variants, or insertions were observed in 21% of the total variants detected (N=25/120). These were detected in hypothetical proteins (N= 12/25, 48%), transposases (N=5/25, 20%), a putative protein YjdJ (N=1/25, 4%), antigen 43 (N=2/25, 8%), cytoskeleton bundling-enhancing antitoxin (N=1/25, 4%), ompF outer membrane porin F (N=1/25, 4%) and D-alanine--D-alanine ligase A (N=1/25, 4%) (**Table S3**). We next tested the hypothesis that the frequency of known AMR genes or virulence factors (VFs) might increase within patients over time, due to exposure to antibiotics and other selective pressures in the critical care hospital environment. Although certain patients (*e.g.* MGH 13) acquired several AMR genes and VFs over time, there was no consistent trend across patients (**Table S3, Figure 5**). Changes were most pronounced in patients MGH 4, MGH 7 and MGH 13, who experienced a putative strain replacement between time points (**Figure 4**, **Figure 5**).

**Figure 5:**
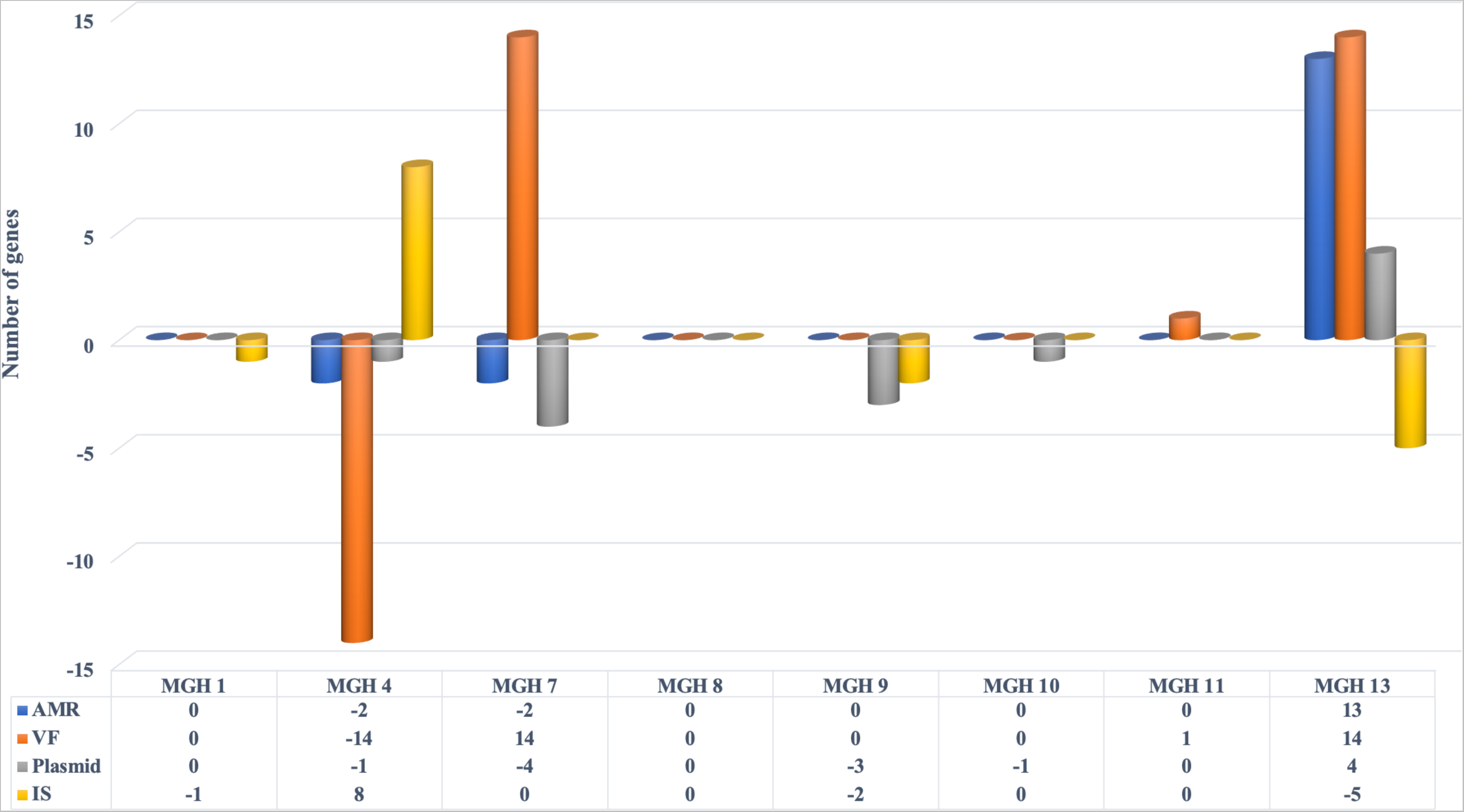
Summary of genes acquired or lost in *E. coli* strains isolated from eight patients at two different points. Gene gains (values above 0) or losses (below 0) over time (between T0 and T1) are indicated for AMR genes, virulence factors (VF), plasmids and insertion sequences (IS).

## Discussion

The carriage rate of resistant *E. coli* isolates has been increasing in both healthcare and community settings. A recent systematic review has shown that at least one in five inpatients worldwide were carriers of ESBL and that the Eastern Mediterranean, which includes Lebanon, had the highest carriage rate (45.6%) (56). Such high prevalence was confirmed in our study while screening patients for resistant Gram-negative bacteria: 70% of patients were colonized with a multi-drug resistant *E. coli* isolate upon admission and during their hospitalization. All patients were critically ill and were given broad-spectrum antibiotics during their hospitalization. Despite the limited number of patients enrolled, we were able to identify that rectal colonization by ST131 subgroup *fim*H30-O25b, clade C1-M27, harboring *bla*_CTX-M-27_ was prevalent, sampled in 33% (N=4/12) of patients. One additional patient harbored a closely related isolate: ST131 subgroup *fim*H41-O25b, clade A-M27, harboring *bla*_CTX-M-27._ Comparative analyses of the five ST131 isolates showed that these strains share an average nucleotide identity of more than 99% and harbor identical virulence factors including type 1 fimbriae (FimH30) and the secreted autotransporter toxin, both of which are involved in uropathogenesis by mediating human bladder epithelial cell adhesion, invasion and biofilm formation (57–60). This strain is also resistant to cephalosporins, carbapenems, quinolones and tetracyclines. ST131 *E. coli* isolates differ from most other MDR *E. coli* by being more pathogenic causing often urinary tract infections (61).

The ST131 strains carrying CTX-M-15 have been reported to cause many infections globally. Recently, the the CTX-M-27-producing clade C1 of *E. coli* ST131 has emerged, and is thought to have epidemic potential both in community and healthcare setting (62–64). To the best of our knowledge, our study reports the first description of *E. coli* ST131 clade C1-M27 circulating in Lebanon. ST131 in Lebanon was first described in 2016 in animals; however, it was associated with *bla*_CTX-M-15_ (39). In our study, we have also shown the dominance of *bla*_CTX-M-15_ in different strains. This gene generally colonizes the gastrointestinal tract of farmed cattle and birds, as well as raw meat intended for human consumption which is a part of the Lebanese weekly diet, and was also identified in surface water (27, 39, 42). Further surveillance should be implemented in Lebanon to understand the transmission of *bla*_CTX-M-15_ and *bla*_CTX-M-27_ from animals to humans. We also identified *bla*_OXA-181_ in the ST131 clone. The *bla*_OXA-181_ gene is mainly found in *E. coli* and *K. pneumoniae*. It was first reported in 2007 (65) and was subsequently identified in several countries (66–69). However, it is not often isolated in ST131 but mostly linked with ST410 and ST1284 (70–72). The presence of both *bla*_OXA-181_ and *bla*_CTX-M27_ genes in ST131 was first observed in a young man with a war-related wound *E. coli* infection, which escalated to a series of recurrent infections over three months; this strain was shown to harbor a *bla*_CTX-M 27_ gene that was transferred from *Morganella morganii* (73). This exemplifies the ability of *E. coli* to rapidly acquire resistance genes from other species and highlights the need for continuous surveillance of gene transfer and resistance evolution in different *E. coli* sequence types.

To date, few studies have investigated within-patient diversity of *E. coli* in the same patient over time (74). In this pilot study, we sequenced *E. coli* genomes upon admission and during hospitalization. A limitation of our study is that we only sequenced two pooled isolates per patient per time point, making it difficult to distinguish strain replacement events from persistent mixed infections. Future efforts should ideally sequence multiple genomes from each sample to better characterize within-patient diversity and improve inference of transmission events (19, 60–62). In addition, using short read sequencing, it was difficult to confidently link resistance genes to plasmids; long-read sequencing could be used to fully assemble plasmids and make these linkages with confidence. Finally, our study only sampled perianal swabs. Sampling additional body sites and tissues could help establish *E. coli* transmission routes within patients.

Despite the limitations, we identified three patients with a possible strain replacement over a few days of hospitalization. In two of these patients sampled at a third time point, the new strain was retained. While we cannot exclude stable co-colonization of these different strains over time, which could have been missed by sequencing two pooled genomes per sample, our study opens the possibility that strain replacement events could plausibly occur during hospitalization. We also hypothesized that the frequency of AMR genes would increase over time in our patients, who were all treated with antibiotics. Although patients and strains both varied widely in their AMR and virulence gene content, there was no evidence for increasing frequency of AMR genes over time. Our work has therefore shown no major changes in SNV occurrence that accounts for disease susceptibility and resistance; this could be due to a lack of power to detect an effect in our small cohort, or could also suggest that resistant strains are already circulating and that resistance is transmitted upon infection rather than evolving within patients.

In conclusion, our results provide evidence for the recent emergence of ST131 subgroup fimH30-O25b, clade C1-M27, in Lebanon and reinforces the need for continuous genomic surveillance of this clone among patients in critical care units. Our study enables longitudinal stain tracking of *E. coli* strains, their colonization dynamics and their diversity within hospitals over time.

## Data Availability

All data produced in the present work are contained in the manuscript

## Supplementary Figures legends

**Figure S1**: **Boxplots displaying the major allele frequency distributions** (i.e., frequency of higher allele, whether alternative or reference) based on single-nucleotide variants and insertions/deletions. These variants were called by Snippy, based on mapped reads to a reference *Escherichia coli* genome. Samples are separated by whether they passed an independent quality control step of whether the whole-genome assembly was >= 8 megabase pairs (Mbp), indicating a potential mixture of strains or other contamination. The number of variants per sample ranges from 8,971-82,636 (mean: 45,920). The box within each boxplots represents the median (centre line) and the 25th and 75th percentiles as the box edges. The whiskers represent 1.5 * inter-quartile range (i.e., the range between the 25th and 75th percentiles), or to the most extreme value. Points indicate outliers outside this range.

**Figure S2: Heatmap summary of Average Nucleotide Identity between each of the 22 strains sequenced in this study.** Values range from 96% ANI to 100% ANI.

**Figure S3: Heatmap showing the plasmid types identified in each of 22 *E. coli* genomes.** The x axis represents the plasmid types.

**Figure S4: Heatmap showing the virulence factors identified in each of 22 *E. coli* genomes.** The x axis represents the virulent genes identified using ABRicate.

## Supplementary Tables legends

**Table S1: Demographic and Clinical characteristics of 14 patients enrolled in the study.** The table includes demographic data in addition to the hospitalization status, risk factors, and antibiotic use during hospitalization. N/A refers to Not Available.

**Table S2: Antimicrobial susceptibility testing results of 32 isolates identified in 15 patients using the disk diffusion method.**

**Table S3: General characteristics of genetic variants identified in seven patients at two different time points.**

