## Supplementary figures and images for "Longitudinal genomic surveillance of multidrug-resistant *Escherichia coli* carriage in critical care patients"

### Supplementary Figure 1

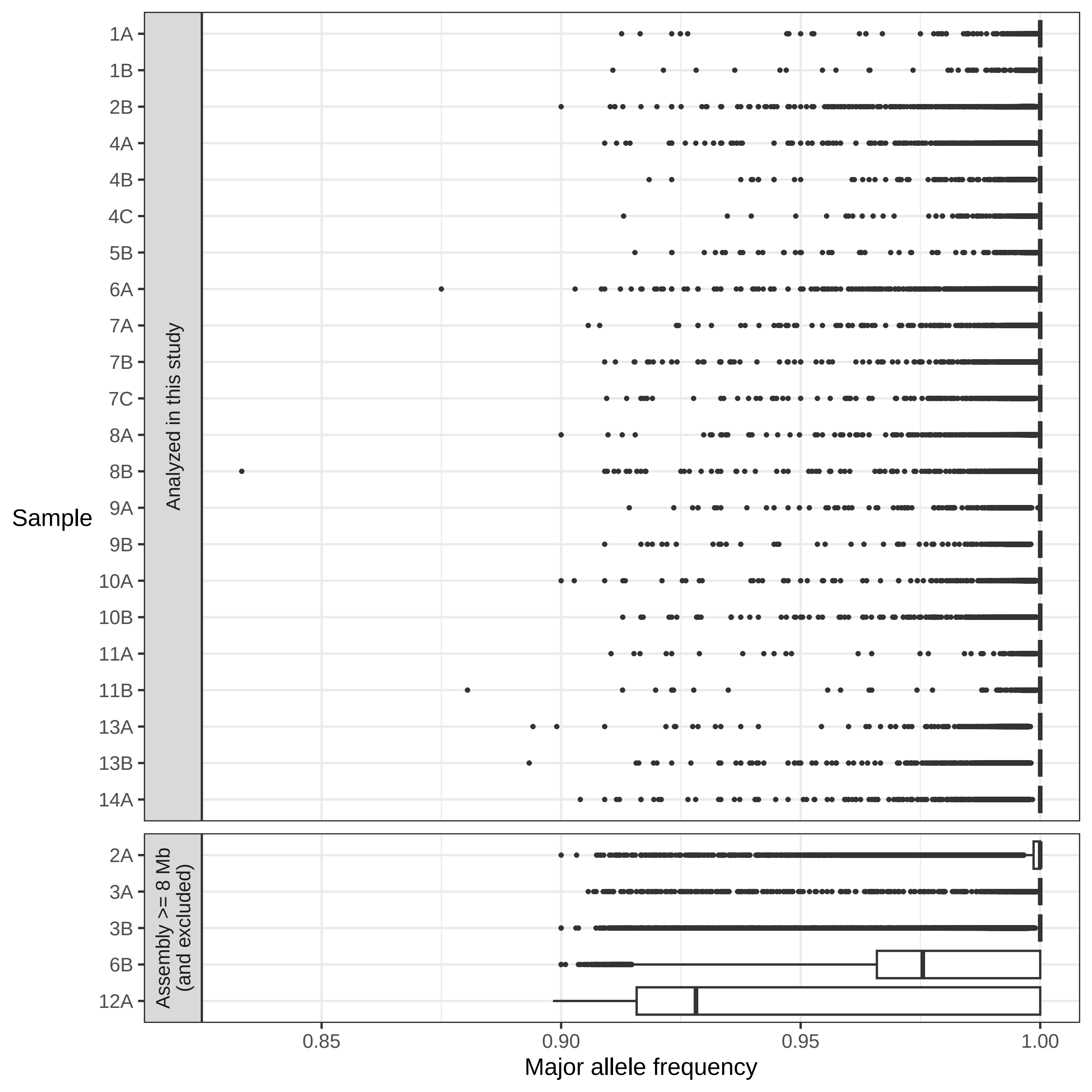

### Supplementary Figure 2

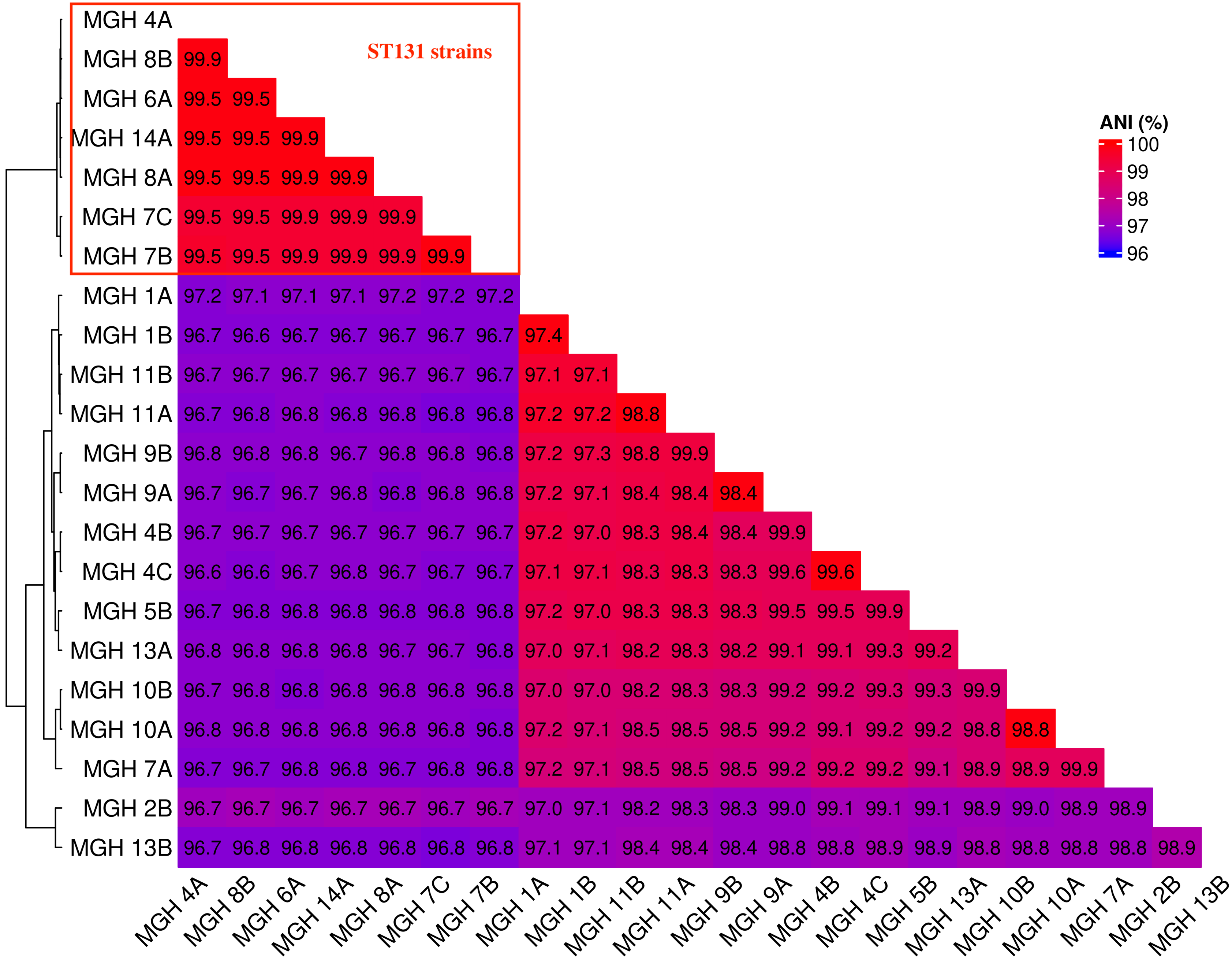

### Supplementary Figure 3

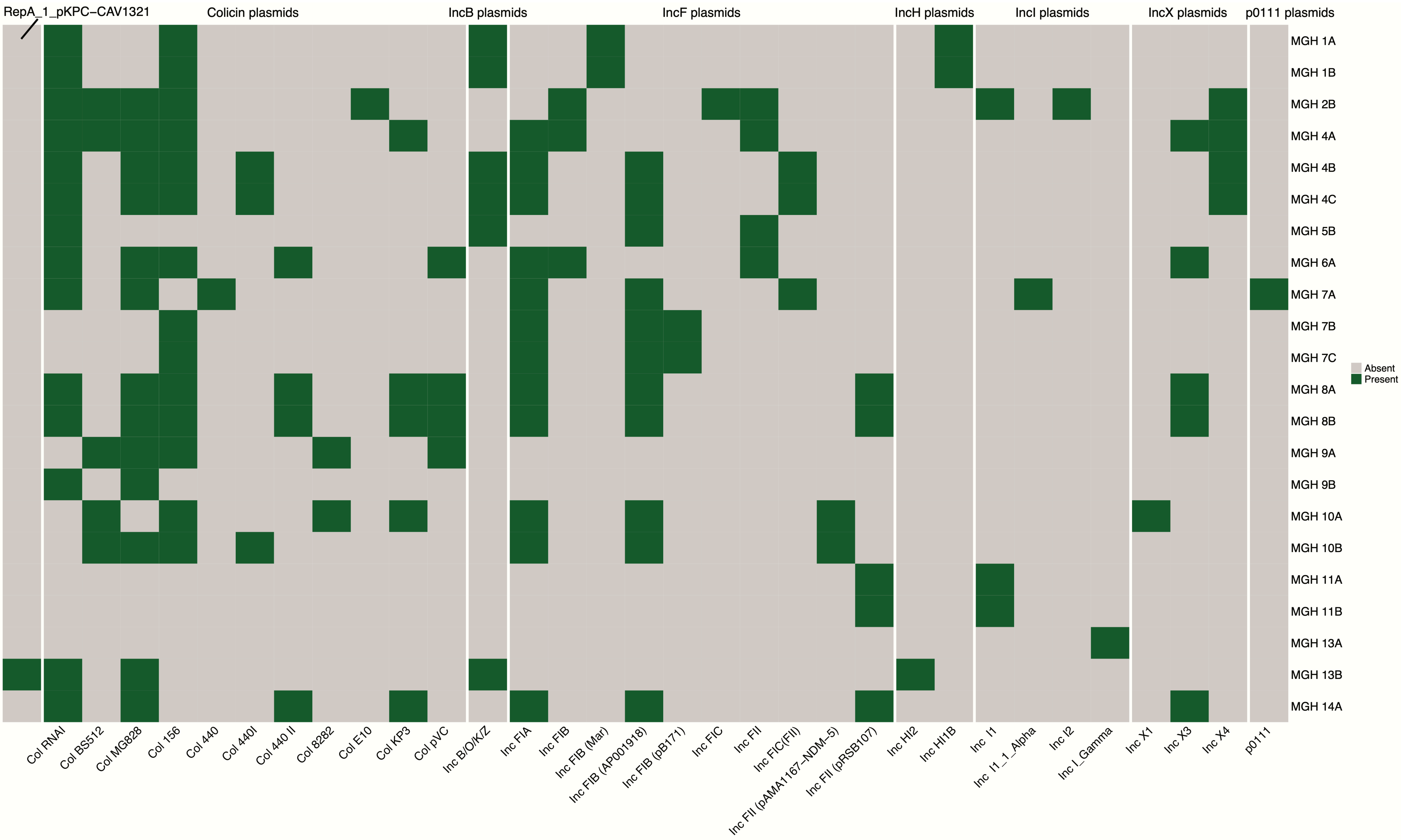

### Supplementary Figure 4

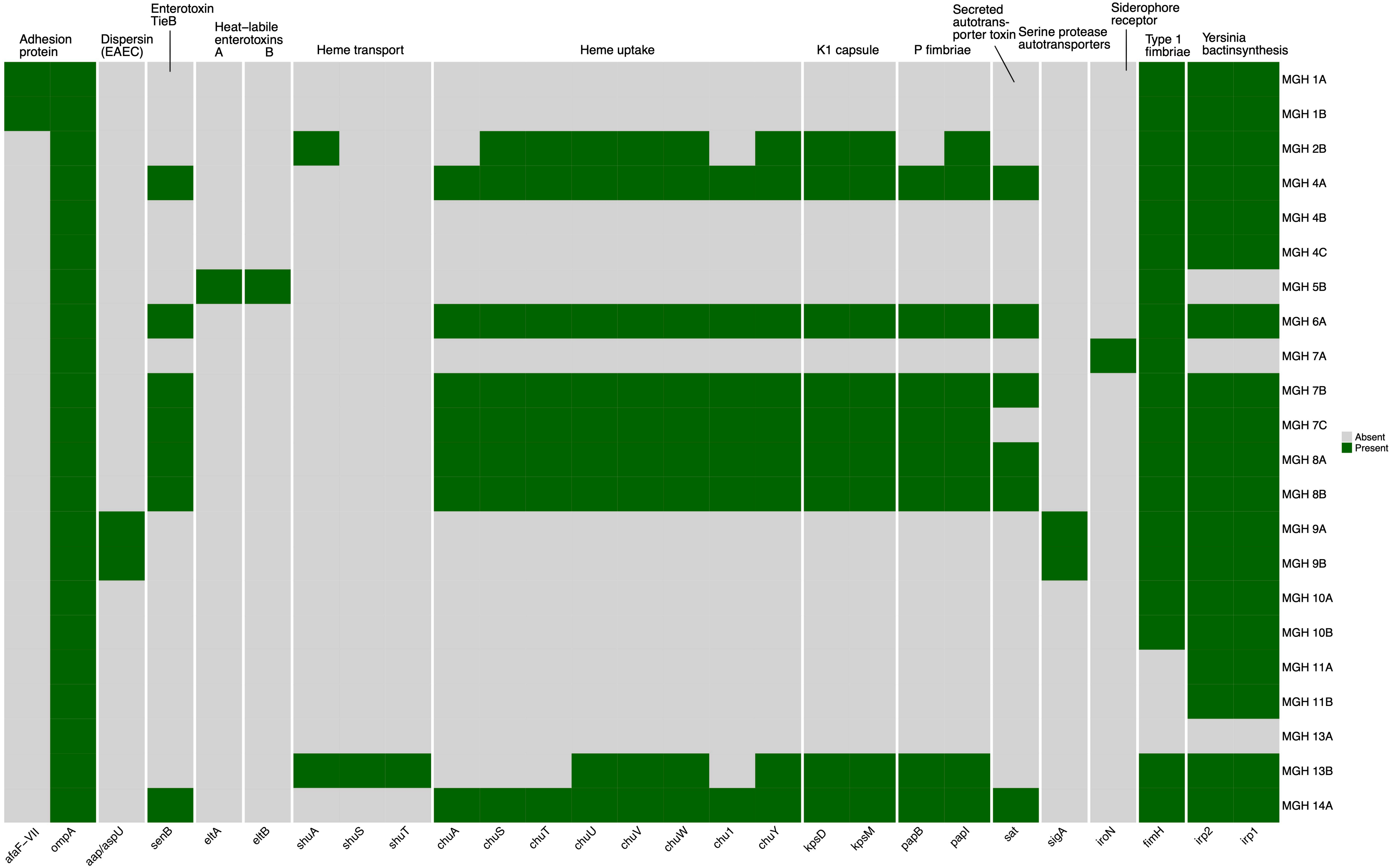
