## Supplementary Table 1 for "Longitudinal genomic surveillance of multidrug-resistant *Escherichia coli* carriage in critical care patients"

| Study ID | Gender | Admission Unit | Cause of admission | Foley catheter | Surgery during hospitalization | Recent antibiotics use (within 3 months) | Antibiotics use during hospitalization | First collection date (T0) | Second collection date (T1) | Third collection date (T2) | Reported infection during hospitalization |
|---|---|---|---|---|---|---|---|---|---|---|---|
| **MGH 1** | F | Hematology/ Oncology | Chemotherapy | None | None | Trimethoprim-sulfamethoxazole (TMP-SMX)<br>Ertapenem<br>Piperacillin/tazobactam<br>Teicoplanin | Meropenem | Nov-21 | Nov-21 | N/A | Urine: *E.coli* (ESBL)<br>Blood: *P.aeruginosa* |
| **MGH 2** | M | ICU | Pneumonia | None | None | N/A | Meropenem<br>Piperacillin/tazobactam<br>Amikacin<br>Vancomycin<br>Levoflaxin | Nov-21 | Dec-21 | N/A | None |
| **MGH 3** | F | Isolation Unit XDR | Pneumonia | Yes | Open cholecystectomy | Ceftriaxone<br>Ceftazidime<br>Ampicillin<br>Amoclan<br>Ceftazidime/Avibactam<br>Vancomycin | Meropenem<br>Ceftazidime/avibactam<br>Amikacin<br>Gentamycin<br>Colistin | Nov-21 | Dec-21 | N/A | None |
| **MGH 4** | M | Hematology/ oncology | Chemotherapy/ Subcranial bleeding | Yes | Craniotomy | N/A | Meropenem<br>Vancomycin<br>Colistin | Sep-21 | Oct-21 | Oct-21 | None |
| **MGH 5** | F | ICU | Pneumonia | Yes | Bedsore | Trimethoprim-sulfamethoxazole (TMP-SMX)<br>Ertapenem<br>Piperacillin/tazobactam<br>Teicoplanin | Meropenem<br>Piperacillin/tazobactam<br>Amikacin<br>Vancomycin<br>Colistin<br>Trimethoprim-sulfamethoxazole | Oct-21 | Oct-21 | N/A | DTA: *A.baumannii* (XDR)-<br>*P. aeruginosa* (XDR)<br>Wound: *A. baumannii* (XDR)-<br>*K pneumoniae* (XDR)- *E.faecium* (VRE)<br>Blood: CoNS |
| **MGH 6** | M | Hematology/ Oncology | Chemotherapy patient with fever | None | None | N/A | Ceftazidime/avibactam<br>Piperacillin/tazobactam<br>Trimethoprim/Sulfamethoxazole | Aug-21 | Sep-21 | N/A | None |
| **MGH 7** | F | ICU | Chemotherapy patient with fever | Yes | None | N/A | Ceftriaxone<br>Ceftazidime/avibactam<br>Meropenem<br>Colistin<br>Levofloxacin | Oct-21 | Oct-21 | Oct-21 | DTA: *E.coli* (XDR)- *S.aurues* (MRSA)-<br>*Candida* species- *Pseudomonas* species<br>Sputum: *S.maltophilia*<br>Blood: *Ecoli* (ESBL)<br>Urine : *Candida* species |
| **MGH 8** | M | ICU | Pneumonia with septic shock | Yes | None | N/A | Meropenem | Sep-21 | Oct-21 | N/A | DTA: *S. aurues* |
| **MGH 9** | F | Isolation Unit XDR | Pneumonia | Yes | None | N/A | Ceftriaxone<br>Piperacillin/ tazobactam<br>Levofloxacin<br>Clarithromycin | Nov-21 | Nov-21 | N/A | None |
| **MGH 10** | F | ICU | Respiratory failure and septic shock | Yes: | Double J insertion | N/A | Meropenem<br>piperacillin/tazobactam<br>Amikacin<br>Vancomycin<br>Colistin | Oct-21 | Nov-21 | N/A | DTA: *P. aeruginosa* (CR) -<br>*A. baumannii* (CRAB) - *C. albicans*<br>Blood: *P. aeruginosa* (XDR)- CoNS |
| **MGH 11** | F | ICU | Pneumonia with septic shock | Yes: | None | N/A | Ceftriaxone<br>Ceftazidime/avibactam<br>Meropenem<br>Colistin<br>Levofloxacin<br>Clarithromycin | Oct-21 | Nov-21 | N/A | Wound: *Candida species* |
| **MGH 12** | M | Isolation Unit XDR | Hemorrhagic shock due to hematuria | Yes | Colostomy | N/A | Meropenem<br>Piperacillin/tazobactam<br>Colistin<br>Amikacin<br>Vancomycin<br>Teicoplanin | Sep-21 | Sep-21 | N/A | DTA: *A.baumannii (* XDR)<br>*P.aeruginosa (* XDR)<br>Blood:*P.aeruginosa*- CoNS<br>Wound: *A. baumannii* (XDR)-<br>*P.aeruginosa* (XDR)-*E. faecium* (VRE)<br>*C. albicans*<br>Urine: *P.aeruginosa* (XDR)-E. faecium |
| **MGH 13** | M | ICU | Esophageal varcies bleeding (liver cirrhosis) | Yes | None | N/A | Ceftriaxone | Sep-21 | Sep-21 | N/A | None |
| **MGH 14** | F | Isolation Unit XDR | Septic arteritis | Yes | None | N/A | Ceftriaxone<br>Piperacillin/ tazobactam<br>Ceftazidime/Avibactam<br>Meropenem<br>Amikacin<br>Vancomycin<br>Teicoplanin | Oct-21 | Oct-21 | N/A | None |
