## Supplementary Table 2 for "Longitudinal genomic surveillance of multidrug-resistant *Escherichia coli* carriage in critical care patients"

| Isolate # | Type of resistant bacteria | Species perianal colonization | Treatment taken during hospitalization | Ampicillin (AMP) | Augmentin (AMC) | Tazocin (TZP) | Cefepime (FEP) | Cefoxitin (FOX ) | Cefotaxime (CTX) | Ceftazidime (CAZ) | Ceftriaxone (CRO) | Cefuroxime (CXM) | Cephalotin (CF) | Ceftazidim/Avibactam (CZA) | Ertapenem (ETP) | Imipenem (IM) | Meropenem (MEM) | Gentamycin (GM) | Amikacin (AN) | Ciprofloxacin (CIP) | Nitrofurantoin (NIT) | Trimethyl sulfate (SXT) | Fosfomycin (FOS) | Tetracyline (TET) | Tigecycline (TGC) | Colistin (CL) | Azithromycin (ATM) |
|---|---|---|---|---|---|---|---|---|---|---|---|---|---|---|---|---|---|---|---|---|---|---|---|---|---|---|---|
| **MGH 1A** | ESBL/CRE | *E..coli* | *Meropenem* | R | R | R | | | R | R | R | R | R | | R | R | R | | | R | | R | | R | | | |
| **MGH 1B** | ESBL/CRE | | | R | R | R | | | R | R | R | R | R | | R | R | R | | | R | | R | | R | | | |
| **MGH 2A** | ESBL/CRE | *E..coli* | *Meropenem- Tazocin - Levoflaxin* | R | R | R | | | R | R | R | R | R | | R | R | R | I | | R | | R | | R | | | |
| **MGH 2B** | ESBL/CRE | | | R | R | R | R | R | R | R | R | R | R | R | R | R | R | | | | | R | | R | | | |
| **MGH 3A** | ESBL/CRE | *E..coli* | *None* | R | R | | R | R | R | R | R | R | R | | R | | | I | | R | | | | R | | | |
| **MGH 3B** | ESBL/CRE | | | R | | | | | R | | R | R | R | | R | | | | | | | R | | R | | | |
| **MGH 4A** | CRE | *E..coli* | *Meropenem- Colistin* | | | | | | | | | | | | R | | | | | | | | | R | | | |
| **MGH 4B** | ESBL | | | R | R | | I | I | R | R | R | R | R | | | | | | | R | | R | | R | | | |
| **MGH 4C** | ESBL | | | R | R | | R | R | R | R | R | R | R | | | | | R | | R | | R | | R | | | |
| **MGH 5A** | CRAB | *A.baumanii* | *Meropenem -Tazocin- Amikacin Colistin-Trimethoprim-sulfamethoxazole* | R | R | | | | R | R | R | R | R | | R | R | R | | | | | R | R | R | | | |
| **MGH 5B** | ESBL | *E..coli* | | | | | | | R | R | R | R | R | | R | R | R | R | | R | | R | | | | | |
| **MGH 6A** | ESBL/CRE | *E..coli* | *Ceftazidime/avibactam- Tazocin* | | | | | | R | R | R | R | R | | R | R | R | | | | | | | R | | | |
| **MGH 6B** | ESBL/CRE | | | R | R | R | R | R | R | R | R | R | R | R | R | R | R | | | R | | R | | R | | | |
| **MGH 7A** | ESBL | *E..coli* | *Ceftriaxone- Ceftazidime/avibactam- Meropenem- Colistin- Levofloxacin* | R | R | | | | R | | | | R | | | | | | | R | | | | R | | | |
| **MGH 7B** | ESBL | | | R | | | | | R | | R | R | R | | | | | | | R | | | | | | | |
| **MGH 7C** | ESBL | | | R | | | | | R | | R | R | R | | | | | | | R | | | | | | | |
| **MGH 8A** | ESBL/CRE | *E..coli* | *Meropenem* | | | | | | R | | R | R | R | | R | | | I | | | | | | R | | I | |
| **MGH 8B** | ESBL/CRE | | | R | R | | I | | R | R | R | R | R | | R | | | I | | R | | R | | R | | I | R |
| **MGH 9A** | ESBL | *E..coli* | *Ceftriaxone- Tazocin- Levofloxacin- klacid* | R | | | | | R | R | R | | R | | | | | | | | | | | R | | | |
| **MGH 9B** | ESBL | | | R | | | | | R | R | R | | R | | | | | | R | | | | | R | | | |
| **MGH 10A** | ESBL/CRE | *E..coli* | *Meropenem- Tazoncin- Amikacin- Colistin* | R | R | R | | | R | | R | | R | | R | R | R | R | | R | | R | | R | | | |
| **MGH 10B** | ESBL/CRE | | | R | R | R | | | R | | R | | R | | R | R | R | R | | R | | R | | R | | | |
| **MGH 11A** | ESBL | *E..coli* | *Ceftriaxone- Ceftazidime/avibactam- Meropenem- Colistin- Levofloxacin- klacid* | R | | | | | R | | | R | R | | | | | | | | | | | | | | |
| **MGH 11B** | ESBL | | | R | | | | | R | | | R | R | | | | | | | | | | | | | | |
| **MGH 12A** | ESBL | *E..coli* | *Meropenem- Piperacillin/tazobactam Colistin-Amikacin- Vancomycin- Teicoplanin* | R | R | R | R | | R | R | R | R | R | | | | | I | | R | | R | | R | | | |
| **MGH 12B** | ESBL | | | R | R | R | R | | R | R | R | R | R | | | | | I | | R | | R | | R | | | |
| **MGH 13A** | ESBL | *E..coli* | *Ceftriaxone* | R | | | | | R | | R | R | R | | | | | | | | | | | | | | |
| **MGH 13B** | ESBL | | | R | R | | | R | R | R | R | R | | | | | | R | | R | | | R | R | | | |
| **MGH 14A** | ESBL | *E..coli* | *Ceftriaxone- Piperacillin/ tazobactam- Ceftazidime/Avibactam- Meropenem- Amikacin- Vancomycin - Teicoplanin* | R | R | | I | | R | | R | R | R | | | | | | | R | | R | | R | | | |
| **MGH 14B** | ESBL | | | R | R | | I | | R | | R | R | R | | | | | | | R | | R | | R | | | |
