## Supplementary Table 3 for "Longitudinal genomic surveillance of multidrug-resistant *Escherichia coli* carriage in critical care patients"

| Patient # | Position | Type of variant | Nucleotide changes | Gene associated with open reading frame |
|---|---|---|---|---|
| **MGH 1** | 133781 | SNV | C=>T | Non-Coding |
| | 643 | SNV | G=>A | Synonymous_variant - Hypothetical protein |
| **Isolate at T1 compared to T0** | 13 | SNV | T=>C | Non-Coding |
| | 30 | SNV | A=>G | Non-Coding |
| | 36 | SNV | A=>G | Non-Coding |
| | 80956 | Complex | CGAT=>TGAC | Synonymous_variant - ISAs1 family transposase ISEc1 |
| | 14629 | SNV | C=>T | Non-Coding |
| | 14666 | SNV | C=>T | Non-Coding |
| | 870 | SNV | A=>T | Non-Coding |
| | 923 | SNV | C=>T | Non-Coding |
| | 1025 | SNV | T=>C | Synonymous_variant - ISAs1 family transposase ISEc1 |
| | 3031 | Complex | CAACT=>AAACC | Missense-variant - ISAs1 family transposase ISEc5 |
| | 48 | SNV | C=>T | Non-Coding |
| | 56 | SNV | T=>C | Non-Coding |
| **MGH 4** | 133573 | SNV | C=>T | Non-Coding |
| | 204751 | INS | A=>ACGGTCAG | Frameshift_variant - ompF Outer membrane porin F |
| **Isolate at T2 compared to T1** | 419 | SNV | T=>A | Non-Coding |
| | 156833 | SNV | T=>G | Non-Coding |
| | 39553 | Complex | ATCG=>GTCA | Synonymous_variant - ISAs1 family transposase ISEc1 |
| | 108554 | SNV | T=>C | Non-Coding |
| | 108560 | SNV | T=>C | Non-Coding |
| | 108577 | SNV | A=>G | Non-Coding |
| | 2120 | SNV | C=>A | Missense_variant - Hypothetical protein |
| | 1257 | SNV | C=>T | Synonymous_variant - Hypothetical protein |
| | 1269 | SNV | C=>T | Synonymous_variant - Hypothetical protein |
| | 1275 | SNV | C=>T | Synonymous_variant - Hypothetical protein |
| | 57894 | SNV | A=>G | Non-Coding |
| | 42632 | Complex | AGTTG=>GGTTT | Missense_variant - ISAs1 family transposase ISEc5 |
| | 343 | SNV | A=>G | Non-Coding |
| **MGH 7** | 59359 | SNV | A=>G | Synonymous_variant - yjdJ_1 putative protein YjdJ |
| | 59368 | SNV | T=>C | Synonymous_variant - yjdJ_1 putative protein YjdJ |
| **Isolate at T2 compared to T1** | 59390 | SNV | T=>G | Missense_variant - yjdJ_1 putative protein YjdJ |
| | 74834 | SNV | G=>A | Synonymous_variant - yjdJ_2 putative protein YjdJ |
| | 4458 | SNV | T=>C | Non-Coding |
| | 10569 | INS | T=>TTGTTTC | Insertion - Hypothetical protein |
| | 2910 | SNV | C=>T | Missense_variant - IS1 family transposase IS1X2 |
| | 333 | SNV | A=>G | Missense_variant - IS3 family transposase IS2 |
| | 415 | SNV | T=>C | Synonymous_variant - IS3 family transposase IS2 |
| | 16368 | SNV | A=>T | Non-Coding |
| | 1385 | SNV | A=>G | Missense_variant - Hypothetical protein |
| | 1436 | SNV | T=>C | Missense_variant - Hypothetical protein |
| | 90 | SNV | C=>T | Non-Coding |
| | 114 | SNV | A=>T | Non-Coding |
| | 135 | Complex | TGGC=>CGGT | Non-Coding |
| | 156 | Complex | AGCT=>GGCA | Non-Coding |
| | 196 | SNV | T=>C | Non-Coding |
| | 252 | SNV | T=>A | Non-Coding |
| | 444 | SNV | A=>G | Synonymous_variant - Hypothetical protein |
| | 469 | SNV | C=>T | Missense_variant - Hypothetical protein |
| | 163 | SNV | T=>C | Synonymous_variant - Hypothetical protein |
| | 752 | SNV | C=>T | Non-Coding |
| **MGH 8** | 2078 | SNV | A=>G | Synonymous_variant - Hypothetical protein |
| | 2156 | SNV | A=>T | Synonymous_variant - Hypothetical protein |
| **Isolate at T1 compared to T0** | 21976 | SNV | G=>A | Missense_variant - Hypothetical protein |
| | 39631 | SNV | G=>A | Synonymous_variant - yjdJ_1 putative protein |
| | 8908 | SNV | G=>C | Missense_variant - Antigen 43 |
| | 9029 | SNV | T=>A | Synonymous_variant - Antigen 43 |
| | 9044 | SNV | G=>T | Synonymous_variant - Antigen 43 |
| | 11429 | SNV | A=>G | Missense_variant - Hypothetical protein |
| | 5424 | SNV | G=>A | Non-Coding |
| | 23877 | SNV | G=>A | Non-Coding |
| | 7024 | SNV | C=>T | Synonymous_variant - Hypothetical protein |
| | 7031 | SNV | T=>C | Missense_variant - Hypothetical protein |
| | 7036 | SNV | A=>G | Synonymous_variant - Hypothetical protein |
| | 7200 | SNV | G=>A | Missense_variant - hypothetical protein |
| | 7258 | SNV | C=>T | Synonymous_variant - Antigen 43 |
| | 7290 | SNV | T=>C | Missense_variant - Antigen 43 |
| | 7297 | SNV | T=>A | Synonymous_variant - Antigen 43 |
| | 4303 | SNV | C=>A | Non-Coding |
| **MGH 9** | 127929 | SNV | C=>T | Missense_variant - Hypothetical protein |
| | 75380 | SNV | T=>C | Non-Coding |
| **Isolate at T1 compared to T0** | 71980 | SNV | T=>G | Non-Coding |
| | 4205 | SNV | T=>G | Synonymous_variant - Hypothetical protein |
| | 21 | SNV | T=>C | Non-Coding |
| | 38 | SNV | A=>G | Non-Coding |
| | 44 | SNV | A=>G | Non-Coding |
| | 18231 | SNV | G=>T | Synonymous_variant - Hypothetical protein |
| | 18240 | SNV | G=>T | Synonymous_variant - Hypothetical protein |
| | 18252 | SNV | G=>A | Synonymous_variant -Hypothetical protein |
| | 12127 | SNV | G=>C | Non-Coding |
| | 1495 | SNV | A=>G | Missense_variant - hypothetical protein |
| | 3758 | SNV | T=>C | Non-Coding |
| | 2948 | Complex | TCG=>CCC | Non-Coding |
| | 30 | SNV | A=>G | Non-Coding |
| | 41 | SNV | T=>C | Non-Coding |
| | 54 | SNV | G=>T | Non-Coding |
| | 64 | Complex | GTC=>TTT | Non-Coding |
| | 899 | SNV | C=>A | Missense_variant - traD_2 Coupling protein |
| | 909 | SNV | G=>A | Missense_variant - traD_2 Coupling protein |
| | 1059 | SNV | C=>T | Non-Coding |
| | 627 | Complex | CTGG=>ATGA | Non-Coding |
| | 642 | SNV | A=>G | Non-Coding |
| | 334 | SNV | A=>G | Non-Coding |
| | 342 | Complex | CGGT=>AGGC | Non-Coding |
| | 264 | SNV | G=>A | Non-Coding |
| **MGH 10** | 15595 | SNV | C=>T | Synonymous_variant - Hypothetical protein |
| | 15610 | Complex | ATCGTCGAGG=>GTCATCCAGC | Synonymous_variant - Hypothetical protein |
| **Isolate at T1 compared to T0** | 15661 | SNV | A=>G | Synonymous_variant - Hypothetical protein |
| | 15685 | SNV | A=>G | Synonymous_variant - Hypothetical protein |
| | 116423 | SNV | A=>G | Non-Coding |
| | 21091 | SNV | C=>T | Synonymous_variant - Small toxic polypeptide LdrD |
| | 21100 | SNV | G=>A | Synonymous_variant - Small toxic polypeptide LdrD |
| | 10454 | SNV | G=>T | Non-Coding |
| | 10524 | SNV | A=>T | Non-Coding |
| | 19817 | SNV | C=>T | Synonymous_variant - DNA primase |
| | 1006 | SNV | T=>C | Missense_variant - Cytoskeleton bundling-enhancing antitoxin |
| | 2014 | SNV | G=>A | Synonymous_variant - Tyrosine recombinase XerC |
| **MGH 11** | 123449 | SNV | G=>A | Missense_variant - D-alanine--D-alanine ligase A |
| | 68938 | Complex | ACAGGT=>TCAGAC | Missense_variant - Hpothetical protein |
| **Isolate at T1 compared to T0** | 69660 | SNV | A=>G | Non-Coding |
| | 69697 | SNV | A=>G | Non-Coding |
| | 1140 | SNV | A=>T | Non-Coding |
| | 1300 | Complex | CTGTG=>GTGTA | Non-Coding |
| | 1316 | SNV | T=>C | Non-Coding |
| | 15488 | SNV | A=>C | Non-Coding |
| | 17654 | SNV | A=>G | Synonymous_variant - Hypothetical protein |
| | 43271 | SNV | T=>C | Non-Coding |
| | 42118 | Complex | AGTTG=>GGTTT | Missense_variant - ISAs1 family transposase ISEc5 |
| | 48 | SNV | C=>T | Non-Coding |
| | 56 | SNV | T=>C | Non-Coding |

NB: The isolates that have > 1000 variants in MGH 4 (T1 versus T0) , MGH7 (T1 versus T0) and MGH 13 (T1 versus T0) were not included in the table since the strains had major differences in the genomic content
SNV is the abbreviation of single nucleotide polymorphism, and INS is the abbraviation of insertion and complex is the abbreviation of combination of SNV
